# Modeling the within-host dynamics of *S. mansoni:* The consequences of treatment frequency and inconsistent efficacy for disease control

**DOI:** 10.64898/2026.02.26.26347231

**Authors:** Larissa C. Anderson, Helen J. Wearing

**Affiliations:** Department of Biology, University of New Mexico, Albuquerque, NM 87131

## Abstract

Schistosomiasis is a neglected parasitic disease caused by various trematode species of the genus *Schistosoma* for which 251 million people needed treatment in 2021. Many mathematical models of *Schistosoma mansoni* transmission incorporate the effect of chemoprophylaxis on parasite burden within the human host. While praziquantel is the most commonly implemented pharmaceutical used to control schistosomiasis, due to its applicability over several species and its negligible side effects, it is not very effective against juvenile schistosomes in humans. This limited efficacy on the juvenile life-stage of the parasite may be an important factor in the persistence of the disease. The demographic consequences of praziquantel use on schistosome population age and sex composition within the human host may obfuscate the effectiveness of these chemoprophylactic control strategies. Furthermore, the effectiveness of this treatment is heavily dependent on the force of infection to humans and the frequency at which these pharmaceuticals are administered. Using a stochastic mechanistic model, we investigated the effects of inconsistent drug efficacy among parasite life stages, varying parasite population structure within the human host, and alternative treatment regimes to the prevailing once-yearly strategy. This allowed us to identify the reduction in infection prevalence under differing infection risk scenarios, parasite population structures at the time of treatment, and treatment schedules. Our results indicate that if elimination is the goal, then widespread (>75% of the population) treatment should be the target and that more frequent treatment schedules are useful up to several treatments a year.

## Introduction

Schistosomiasis is a neglected parasitic disease caused by various trematode species of the genus Schistosoma. Chronic schistosomiasis infections are of concern as they can lead to serious complications such as enlargement of the liver and spleen, and kidney damage [1]. Despite consistent control efforts, this disease is endemic in 51 countries, with 90 percent of cases occurring in sub-Saharan Africa as of 2021 when an estimated 251.4 million people needed treatment for this infection, and an estimated 1.7 million DALYs were lost [2,3]. The most common of the schistosoma species in sub-Saharan Africa, *Schistosoma mansoni*, uses humans and planorbid snails as obligate hosts, providing multiple avenues for parasite control. Current control strategies include improvement in water availability and sanitation infrastructure, education, molluscicide use on snail populations, and mass drug administration (MDA) in the human population. While modeling efforts have indicated that repeated MDA has resulted in reduction in morbidity since the year 2000, elimination of *S. mansoni* in sub-Saharan Africa has proven elusive [4]. As a result, schistosomiasis is one of the neglected tropical diseases identified by the World Health Organization (WHO) for elimination as a public health problem by 2030, a target that will be achieved when <1% of school-aged children have high intensity infections [5]. In order to achieve these elimination goals, the WHO has called for the incorporation of models to assist in local strategy for elimination [6].

Mass drug administration is the primary form of control and as such we propose a model to understand the robustness of the Schistosoma transmission system to various MDA regimes. Recent stochastic modeling efforts have shown that elimination may only be possible in highly endemic areas if the proportion of individuals who are never treated is extremely low and overall population coverage remains high, and that strategies more frequent than once yearly MDA would be needed to reach elimination in high intensity areas[7]. Also, while less frequent MDA strategies may be sufficient to achieve elimination as a public health problem in moderate prevalence environments, in high prevalence areas current MDA strategies could lead to rebound in prevalence [7–10].

In contrast to almost all prior studies that evaluated the impact of MDA on human populations, we focus on the effectiveness of MDA within each individual human host and how that translates into population-level outcomes. A recent study has also included the impacts of MDA on stratified parasite populations within individual hosts, however they primarily investigated theoretical future drug efficacies on these parasite populations, while we focus only on currently available pharmaceutical options[11]. Our inclusion of individual-level dynamics during MDA allows a unique insight towards areas with persistent transmission despite consistent MDA and to identify avenues that can maximize control efforts and help reach the morbidity reduction and elimination goals set by the WHO [4,12].

Within the definitive human host, *S. mansoni* goes through several developmental stages which are differentially impacted by current drug treatments. After initial penetration of the skin, Schistosoma cercariae migrate in the bloodstream through the lungs to the liver. The cercariae develop into schistosomulae and these mature into adults in approximately four weeks. After maturation, the adult schistosomes migrate to the mesenteric vein and mate. The female worm lodges itself into the groove of the male schistosome and they remain mated, producing hundreds of eggs per day for their lifetime [1,13,14]. Adult schistosome worms can then live for up to 30 years, though on average they survive for 3-5 years [1,13,14]. In general, there is a large sex bias among adult worms - usually a 2.5:1 male to female ratio – which results in a high probability that a female worm is mated even in low adult worm population densities [15]. Together these life-cycle characteristics result in a robust transmission system.

Currently, the anti-helminth drug, praziquantel, is the most heavily utilized treatment worldwide for schistosomiasis [4,5]. Praziquantel has been in widespread use since 1984 as a first-line treatment choice due to its low cost, minimal side effects and efficacy against all human schistosomiasis species [16]. It has been used extensively in large-scale yearly preventative chemotherapy campaigns in the last decade to reduce overall morbidity and chronic complications. However, praziquantel is not without issue: it is not very effective against juvenile worms, particularly those aged 1-4 weeks [16]. While there are various theories as to the cause of reduced efficacy in juvenile worms there are currently no definitive studies. Cure rates for *S*.*mansoni* infection with praziquantel have been variable, often between 30 and 100%, though it is usually above 72% [16,17]. Overall, while praziquantel has proven cost effective and efficacious in reducing morbidity at the individual level, repeated MDA campaigns among school-aged children in endemic regions do not often result in elimination at the population level [18].

Multiple modeling studies have focused on the prospect of elimination and morbidity control through MDA with praziquantel [19–22]. While some have indicated that a praziquantel based intervention has inadequacies they have conflicting conclusions on whether the current MDA strategies are sufficient for morbidity reduction and elimination despite the persistence of “hotspots” and areas recalcitrant to repeated MDA campaigns [20–26]. However, these models have not included the differential efficacy of praziquantel among schistosome life stages, and the majority do not explore MDA at frequencies of more than once a year for lower prevalence settings, and twice a year for high prevalence settings.

We developed a stochastic mechanistic mathematical model that includes the differential life-stage mortality and numerous MDA treatment schedules to assess the impact these factors have on the success of MDA in achieving morbidity reduction and elimination targets. This model estimates that if elimination is the target, then once yearly MDA is not sufficient except in extremely low burden areas where there is broad (>75%) treatment coverage for the population contributing to transmission. Furthermore, in moderate and high burden areas, to achieve a large reduction in burden there should be multiple praziquantel treatments a year and (>50%) treatment coverage for the population contributing to transmission.

## Methods

### Model Structure

#### Schistosome Population

To address the potential impact of various MDA strategies, we developed a continuous-time Markov chain model, simulated using the tau-leap method. The use of stochastic processes and integer-valued variables allows for schistosome population die-out and therefore local elimination. The model framework includes nine different stages of schistosoma development within an individual human host, which correspond to differential susceptibility to praziquantel treatment and the length of time spent in each developmental stage (Figure 1, Table 1) [27]. This includes declining efficacy of praziquantel for juvenile stages and a lower efficacy in adult male schistosomes (Table 2).

**Figure 1.**
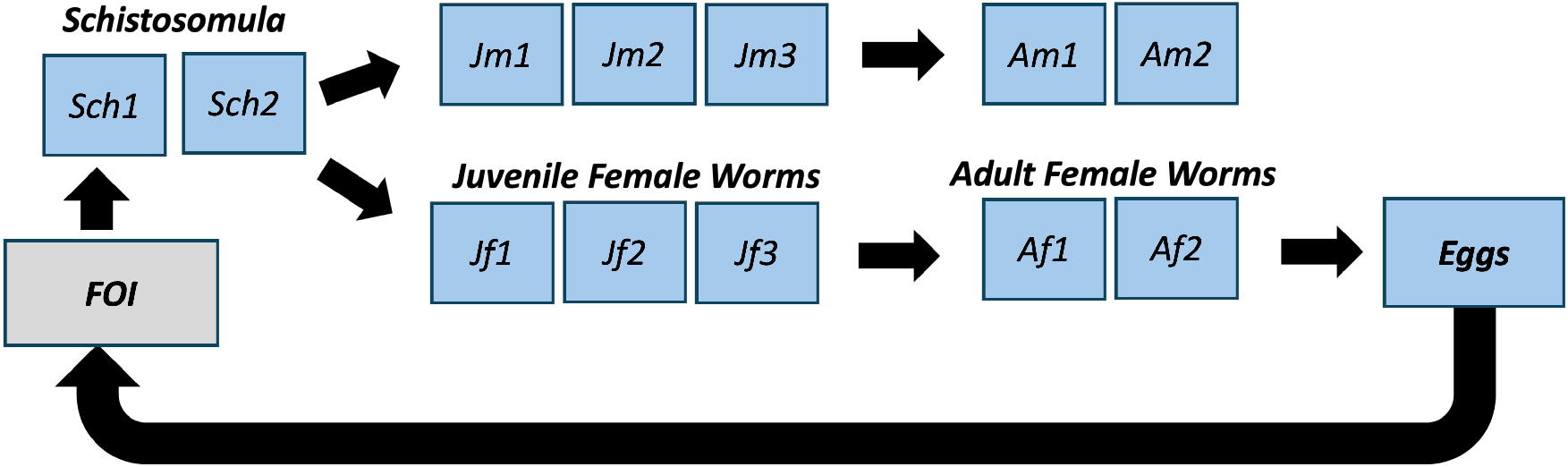
Conceptual diagram of model structure. The parasite population contains three life stages: schistosomulae, juvenile and adult. Within these life stages there are subdivisions corresponding to the sex of the parasite and days spent within each life stage which correspond to differential drug efficacy.

**Table 1:** Description of model variables.

| Variable | Description |
| --- | --- |
| *Sch<sub>1</sub>* | Schistosomulae – Aged 0-7 days |
| *Sch<sub>2</sub>* | Schistosomulae – Aged 8-14 days |
| *Jf<sub>1</sub>* | Juvenile female worms – Aged 15-21 days |
| $Jf_2$ | Juvenile female worms – Aged 22-28 days |
| $Jf_3$ | Juvenile female worms – Aged 29-35 days |
| $Jm_1$ | Juvenile male worms – Aged 15-21 days |
| $Jm_2$ | Juvenile male worms – Aged 22-28 days |
| $Jm_3$ | Juvenile male worms – Aged 29-35 days |
| $Af_1$ | Adult female worms – Aged 36-42 days |
| $Am_1$ | Adult male worms – Aged 36-42 days |
| $Af_2$ | Adult female worms – Aged 43+ days |
| $Am_2$ | Adult male worms – Aged 43+ days |
| $E$ | Eggs |
| $B$ | Force of infection to humans |

**Table 2:** Description of model parameters.

| Variable | Description | Value | Source |
| --- | --- | --- | --- |
| $A_1$ | Rate of transition from schistosomulae to juvenile worms | 1/7 per day | [26] |
| $A_2$ | Rate of transition of juvenile worms to adult worms | 1/7 per day | [26] |
| $U_1$ | Mortality rate for schistosomulae | 1/1095 per day | [13] |
| $U_2$ | Mortality rate for juvenile worms | 1/1095 per day | [13] |
| $U_3$ | Mortality rate for adult male worms | 1/1095 per day | [13] |
| $U_4$ | Mortality rate for adult female worms | 1/1095 per day | [13] |
| $P_1$ | Proportion of new worms that are male | .72 | [14] |
| $P_2$ | Proportion of new worms that are female | .28 | [14] |
| $R$ | Egg production per mated adult female worm | 350 per day | [12] |
| $V$ | Probability that a female worm is mated | .95 | [27] |
| $Y$ | Proportion of eggs that pass out of human host | .5 | Estimated |
| $Tx_1$ | Efficacy of Praziquantel on Sch<sub>1</sub> | 85% | [26] |
| $Tx_2$ | Efficacy of Praziquantel on Sch<sub>2</sub> | 70% | [26] |
| $Tx_3$ | Efficacy of Praziquantel on Jm<sub>1</sub> | 50% | [26] |
| $Tx_4$ | Efficacy of Praziquantel on Jm<sub>2</sub> | 35% | [26] |
| $Tx_5$ | Efficacy of Praziquantel on Jm<sub>3</sub> | 15% | [26] |
| $Tx_6$ | Efficacy of Praziquantel on Jf<sub>1</sub> | 50% | [26] |
| $Tx_7$ | Efficacy of Praziquantel on Jf<sub>2</sub> | 35% | [26] |
| $Tx_8$ | Efficacy of Praziquantel on Jf<sub>3</sub> | 15% | [26] |
| $Tx_9$ | Efficacy of Praziquantel on Af<sub>1</sub> | 95% | [26] |
| $Tx_{10}$ | Efficacy of Praziquantel on $Af_2$ | 95% | [26] |
| $Tx_{11}$ | Efficacy of Praziquantel on $Am_1$ | 60% | [26] |
| $Tx_{12}$ | Efficacy of Praziquantel on $Am_2$ | 60% | [26] |
| $Sc$ | Scaling parameter | 1/500000 | Estimated |

#### Host Population

The human population was kept constant at a size of 100 individuals to be representative of a small community. The initial worm burden profile for the population was set to represent the WHO classes of either: low (<10%), moderate (10-50%) or high (>50%) infection prevalence. Within the infected human population, the worm burden varied for each individual. We calculated the number of mated worm pairs necessary to produce the eggs per gram of feces, (<100, 100-399, >400) which correspond to the WHO classification of low, moderate or heavy burden. The proportion of infected individuals in each burden class was set and an individual human’s schistosome population was then drawn from a distribution of mated worm pairs corresponding to the burden level. The model was run with various combinations of initial population burden (<30%, 50%, >75%), and proportions of individual infection intensity for the infected portion of the human population.

#### MDA Intervention Scenarios

We employ a nested design to isolate the impact of the following: population and individual parasite burden; assumptions about feedback on the force of infection with treatment; MDA campaign length; and MDA frequency. The stochastic model was run for intervention scenarios of 1-6 years in length. The praziquantel MDA was applied to each individual in the population and the schistosomes within each individual decreased in accordance with praziquantel efficacy for each life stage (Table 2). Praziquantel treatment was applied at intervals varying in length from 1-12 months. The treatment was either applied at the same frequency for the duration of the intervention time, for example every two months for 5 years, or it was applied at the same frequency for a set number of doses, for example three treatments, each 2 months apart.

#### Force of Infection Scenarios

The force of infection (FOI) experienced by each individual human is modeled in three different ways: (i) unrelated to egg production; (ii) linked at various proportions to the overall population egg production (50%,75%, 100%); or (iii) linked to both the total population egg production and the individual’s initial egg burden. All corresponding model equations are presented in the supplemental information.

The case of unlinked FOI (scenario i) serves as a proxy for treatment of a subset of the population which does not impact the miracidial population and therefore the FOI from snails to humans. We consider this as a null case where there is no feedback and population-wide downstream effects from our MDA interventions. If the FOI is set to be linked to the egg production of the entire population (scenario ii) then the population FOI is scaled to the mean schistosome egg population. Consequently, there is feedback between egg production and the risk of infection. It is important to note in the population-linked FOI scenarios every individual in the population experiences the same FOI regardless of their initial worm burden. When the FOI is linked to the egg production of the total population and the initial egg production of the individual (scenario iii), the initial egg burden is a measure of an individual’s initial intensity of infection. In this scenario the initial infection intensity is assumed to be influenced by the individual’s risk of infection, either due to susceptibility to the parasite or due to behaviors contributing to exposure. These factors are not considered to change over the course of the intervention, therefore a human with a higher-than-average burden prior to treatment will have a higher FOI than an individual with an initial worm burden equal to the mean population worm burden. The effect of the mean population egg production on the FOI was modified by a scaling parameter. This scaling parameter was included to incorporate the indirect impact of egg burden on the force of infection. This estimated parameter was used to adjust the total number of eggs produced, as a proxy for the large dilution of the impact of schistosome eggs which occurs after they are released into the environment and the asexual parasite reproduction occurring in the snail host segment of the schistosome life cycle. Sensitivity analyses of this scaling parameter were performed and are presented in SI Figure 23.

#### Outcome metrics

The metrics we used to evaluate the efficacy of MDA treatment timings include mean worm burden, adult female schistosome population size, total egg population size, the proportion of individuals who experience total worm die out, adult female schistosome die out, the proportion of time during the intervention period with zero egg production and the schistosome population two years after MDA cessation. The mean worm burden is the standard metric used for describing individual schistosomiasis burden. The adult female schistosome population size is important as it correlates to egg production, which is the primary source of morbidity in individuals. Total egg population size is a potential indicator of the impact that the population may have on ongoing transmission, by introducing these eggs into the environment. The proportion of individuals experiencing total worm die out is a measure of elimination at an individual level. Transmission interruption is indicated by both adult female schistosome die out and the proportion of the intervention period with zero egg production. To establish confidence in the simulation results, all stochastic scenarios were run for 1,000 instances with identical initial conditions. The rebound of the schistosome population after intervention cessation was also evaluated under various FOI values.

#### Stage Dependent Efficacy

To establish the impact of stage-dependent mortality on the treatment outcomes, a “control” baseline model wherein the entire parasite population was assumed to experience either a very low (70%) or more standard (90%) reduction upon treatment was used for comparison (SI Figs 2-3). Oxamniquine, another treatment option for *S. mansoni*, with differing schistosome stage and sex-specific efficacy was also evaluated (Supplemental Figs. 4-5).

## Results

### MDA Intervention Scenarios: Intervention Length

To evaluate the impact of repeated MDA on a representative human population in an endemic setting, praziquantel treatment was applied at varying intervals over intervention lengths of 2, 3, and 5 years. As routine MDA is recommended for environments with endemic schistosomiasis, the initial prevalence in the human population was set above 50%, though simulations with a low initial population burden were run for comparison (SI Figs. 6-13). An example of an initial schistosome population is represented in Figure 2a. This population prior to praziquantel treatment has high levels of prevalence and, consistent with field studies, the majority of high burden individual infections clumped in a small proportion of the population [28,29]. The worm burden in a population prior to treatment at various initial FOI (B) levels is shown in SI Fig 1 and demonstrates the skewed distribution common in field studies. A single iteration of the alteration of the schistosome burden in individuals post treatment is shown in Figure 2b, with treatment applied at six-month intervals over a two-year intervention. In the Figure 2b scenario there is a marked reduction in schistosomes per individual after treatment, with some rebound in schistosome population between the 6-month treatment intervals. As praziquantel is overall quite effective this contraction in the schistosome population is typical directly following treatment.

**Figure 2.**
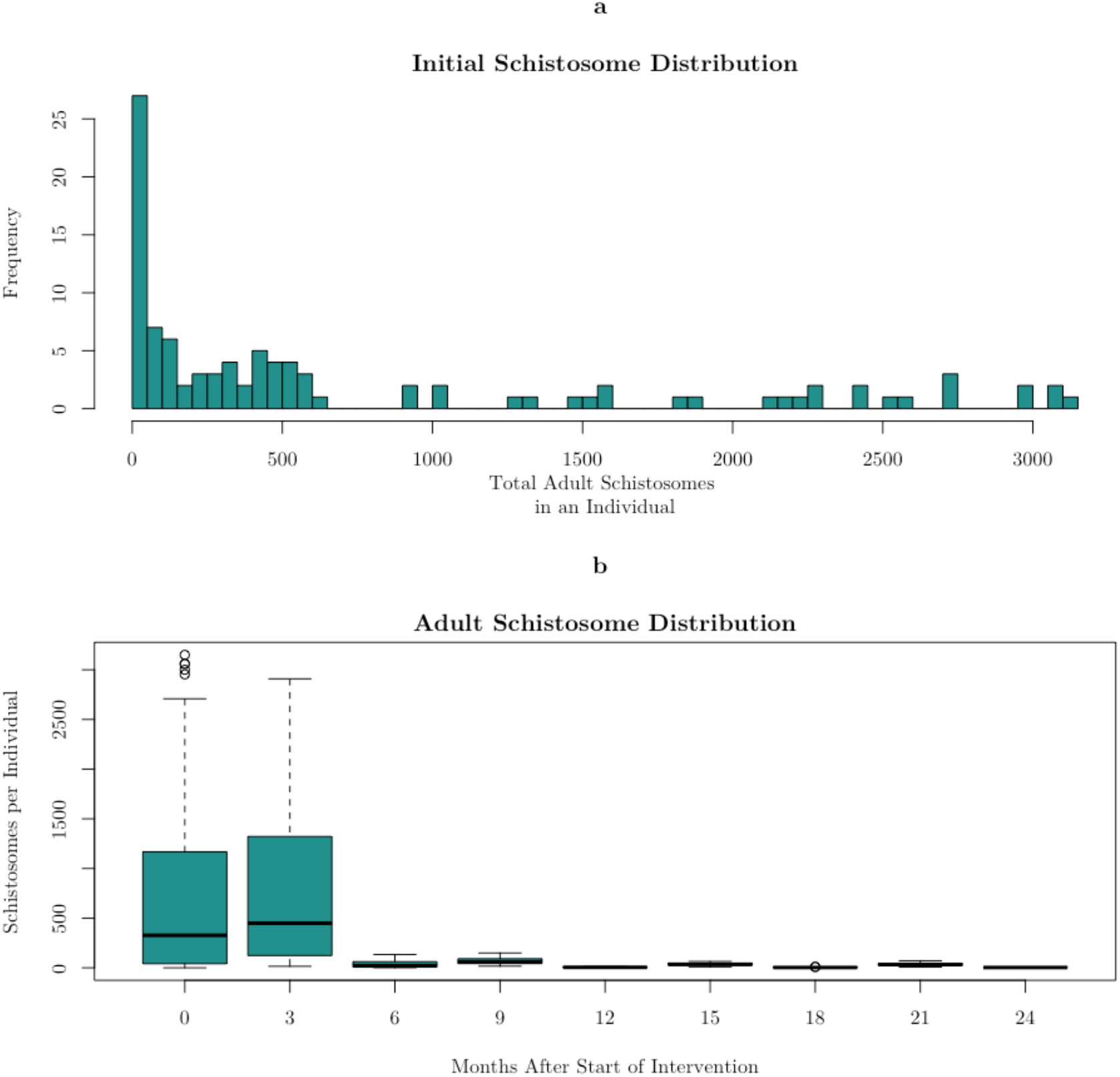
A representative population distribution of adult worms in a moderate risk human population. **Panel a:** Initial adult worm population prior to treatment in a population of 100 individuals. **Panel b:** Adult worm population over a 2-year intervention period with praziquantel treatment applied every 6 months.

### MDA Intervention Scenarios: FOI Assumptions

While there are large reductions in the overall schistosome population at both the individual and community level after treatment, the longer-term effectiveness of MDA varies considerably depending on the initial FOI, the frequency of treatments and the extent to which FOI is linked to the egg production in the treated population (Figure 3). Under the low initial FOI scenario, (B=.5), even without any feedback, and therefore reduction on the FOI after MDA, the mean worm burden is very low after 3 years of consistent treatment. With the linked FOI scenarios, where the FOI over the course of the intervention is impacted by the reduction in eggs produced by the population, there is negative feedback and the result is lower mean worm burden. This diminishment of the FOI and the subsequent effect on schistosome population size is more pronounced with increasing linkage of FOI to the mean number of eggs excreted into the environment. In the model with 100% of the FOI modified by the mean number of eggs, it is assumed that the entirety of the human population contributing to the FOI is included in the MDA treatments. With 100% of the population treated with praziquantel over 3 years, once-yearly treatment is sufficient for elimination. However, the 50% and 75% partial FOI linkage scenarios display mean worm burden values intermediate to the unlinked and 100% linked circumstances. Therefore, mean worm burden increases as a product of increasing length of time between treatments (Figure 3). At higher initial FOI values (B=2-10), low mean worm burden is only achieved through high levels of population treatment, or MDA treatment schedules that are more frequent than the standard once yearly administration. At very high initial FOI values (B=25), low mean worm burdens are only possible with the majority of individuals treated and frequent MDA.

**Figure 3.**
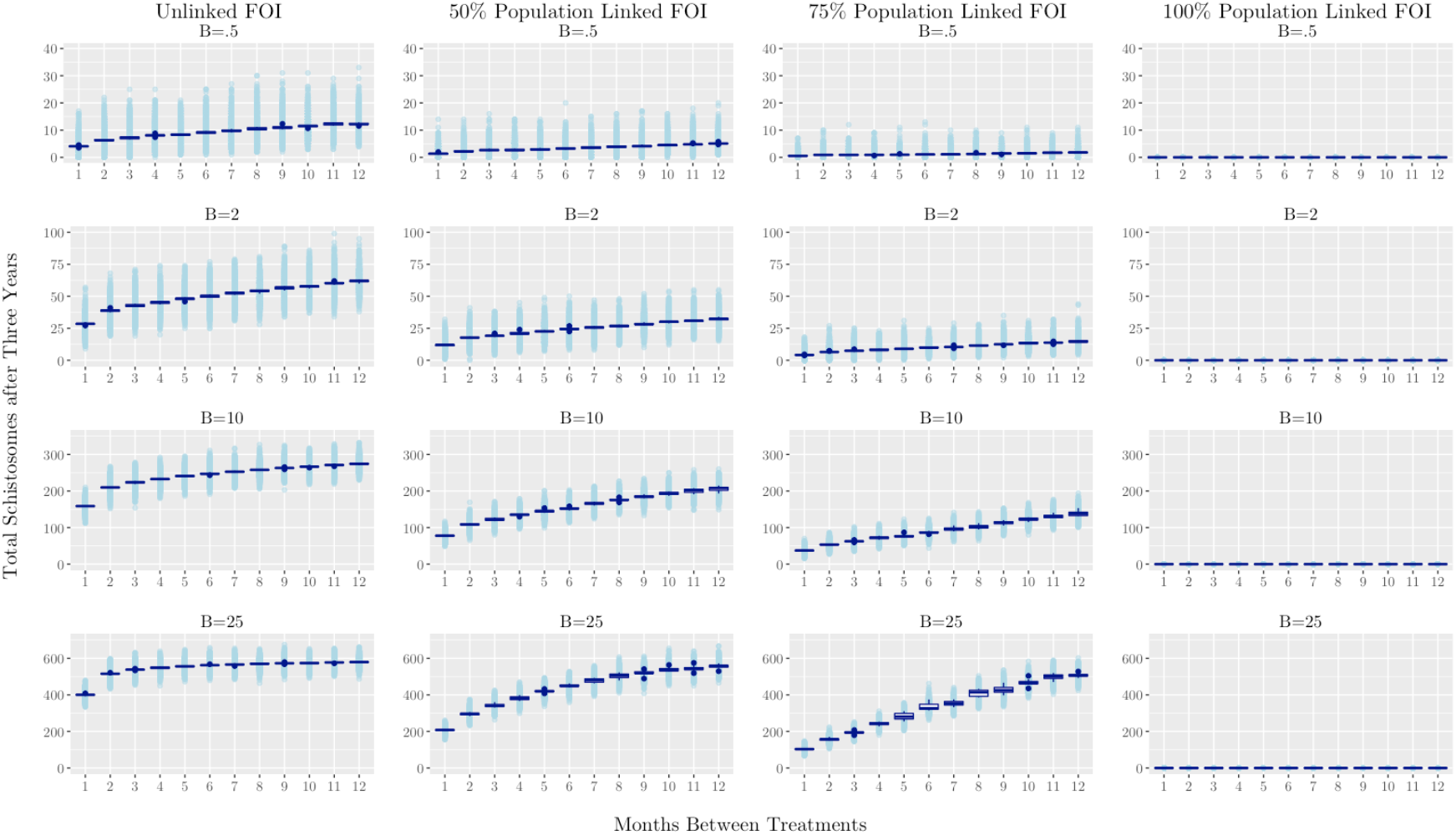
Total worm burden after a 3-year intervention period under varying treatment timing regimes. As the initial FOI (B) increases from.5-25, the total schistosomes after the intervention increases (top to bottom in the panels). As the linkage of FOI to the population egg production increases from 0 (scenario i) to 50, 75 and 100% (scenario ii) the total number of schistosomes tends to decrease (left to right in the panels). The boxplots show the mean adult worm burden for the population over 1,000 runs, while the shaded points show the adult worm burden in each individual in the population over 1,000 runs.

Adult female population size is correlated with overall egg production, which is the primary source of morbidity in humans and is essential to continued transmission. Similar to the total schistosome population, the adult female schistosome population also exhibited persistence under situations with yearly MDA, moderate or high initial FOI and treatment coverage of 75% or less (Figure 4). If there is not 100% linked FOI, then only under a very low FOI regime (B=.5) or moderate FOI (B=2) and very high coverage (>75%) is it possible to achieve adult female population sizes per human of less than 5 under a once yearly MDA schedule (Figure 4). Low adult female populations indicate the possibility of breaks in transmission competency. The egg production of the population mirrored the adult female population, though elimination was not achieved by the end of the intervention period in high FOI situations with yearly MDA (SI Figs. 10-11).

**Figure 4.**
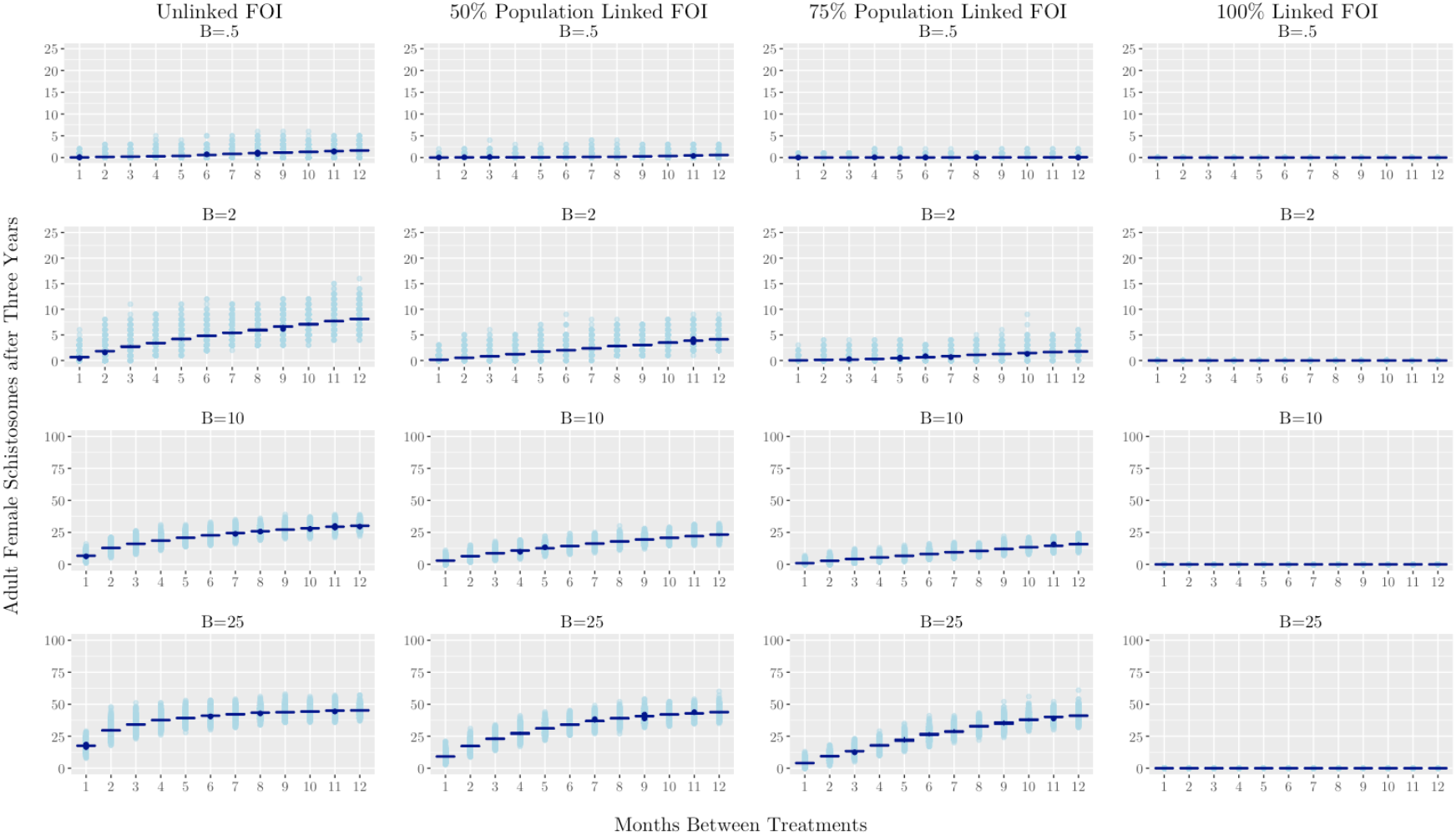
Adult female schistosomes after a 3-year intervention period under varying treatment timing regimes and FOI initial values (B) and feedback types. Die out or very low population size (<5) of adult females with yearly MDA is only achieved, under high functional MDA coverage (75-100% linked FOI). The boxplots show the adult females in the entire human population over 1,000 runs. The adult females for each individual human are represented by the shaded points.

For all initial FOI values, the FOI that is linked to each individual’s egg burden displays qualitatively identical results as the 100% population linked FOI (SI Figs. 6-10). Intervention lengths longer than 3 years showed behavior similar to the 3-year length, indicating a stagnation in effectiveness with longer intervention lengths if FOI is relatively consistent and treatments were given at similar intervals and population coverage (SI Figs. 8-9). Diminishing returns on intervention lengths greater than 3 years indicate that it may be beneficial for the allocation of resources to center on more frequent treatments and greater treatment coverage rather than solely increasing MDA campaign length.

### MDA Intervention Scenarios: Number of Treatments

We also explore a second option for evaluating intervention effectiveness. Rather than a set goal of elimination or specific reduction in burden within a certain time frame, we also simulate outcomes after a set number of treatments. This may be useful in situations where there is a limitation on doses but the logistical ability to administer these treatments at a variety of time points. Lower schistosome populations are seen if the treatments are given at shorter intervals, particularly one month apart (Figure 5). However, under very low FOI conditions, the impact of treatment timing was minimal, instead the targeted treatment of the portion of the population contributing to the FOI was a better indicator of success in achieving elimination (Figure 5). Under moderate and high initial FOI circumstances, treatments applied in quick succession led to a greater reduction in total schistosome population than treatment intervals closer to a year. Elimination was only achieved in moderate or high FOI situations when there was very high treatment coverage (Figure 5). There is not an increase in the effectiveness of the MDA with an increased number of treatments applied at the same interval (SI Figs. 14-17). Consequently, after three treatments there are diminished returns. Therefore, in a scenario with a limited number of doses available, a greater impact on the mean worm burden would be achieved if doses were administered to more individuals in the population for up to three treatments rather than extending the length of the intervention. Treatment regimes that only included two treatments were not as effective as three or more treatments (SI Figs. 12-13,16-17). The adult female and egg populations followed the same patterns exhibited by the mean worm burden (SI Fig 12-17).

**Figure 5.**
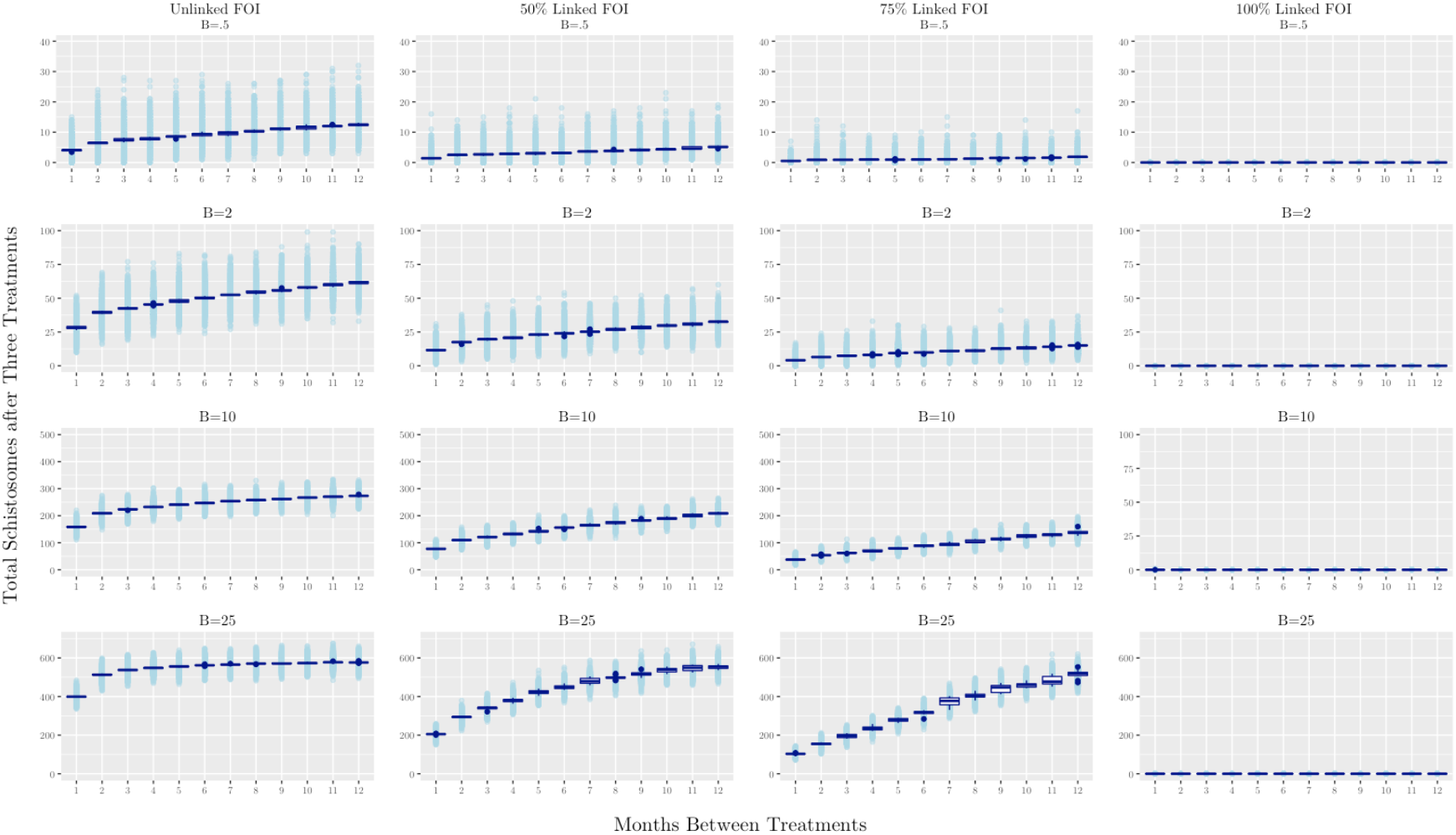
Total worm burden after three treatments under various FOI initial conditions (B), types and treatment intervals. The boxplots show the mean adult worm burden for the population over 1,000 runs, while the shaded points show the adult worm burden in each individual in the population over 1,000 runs. Under low FOI conditions more frequent treatment did result in lower total schistosomes versus once yearly treatment, but the proportion treated (50, 75, 100% linked FOI) had a greater impact on the overall levels than treatment timing. Elimination was only achieved in moderate or high FOI situations when there was very high treatment coverage (100% linked FOI).

### Transmission Interruption

We evaluate two measures of transmission interruption, the proportion of individuals who experience total die out of adult females at any point during the intervention, and the proportion of time where zero eggs are produced by the population. The proportion of individuals who experience die out of adult females are therefore not producing eggs which result in individual morbidity as well as potentially contributing to transmission. Whereas the proportion of time where zero eggs are produced by the population indicates a break in the transmission cycle. The overall adult female population size at the end of 2–5-year intervention period is similar regardless of the initial burden profile of the population (Figure 4 & SI Figs 7,9). However, the proportion of individuals who experience a total die out of adult females at some point during the intervention is greater in those populations with a low initial worm burden (Figure 6 & SI Figs. 15,18). With a low initial worm burden there are more frequent, but very transient, die out events. In moderate or high FOI settings, there are thresholds, between 1–3-month treatment intervals, where there is a transition from a high proportion of individuals experiencing a die out of adult females to a much lower proportion (Figure 6). It should also be noted that in certain instances, after 10-or 11-month treatment intervals there is a lower die out than at 12 months. This disparity reflects the same total treatments for 10–12-month intervals within the 3-year intervention but the greater time for rebound in the schistosome population after the last treatment for the shorter time intervals (Figure 6, SI Table 2, SI Figs. 6-7).

**Figure 6.**
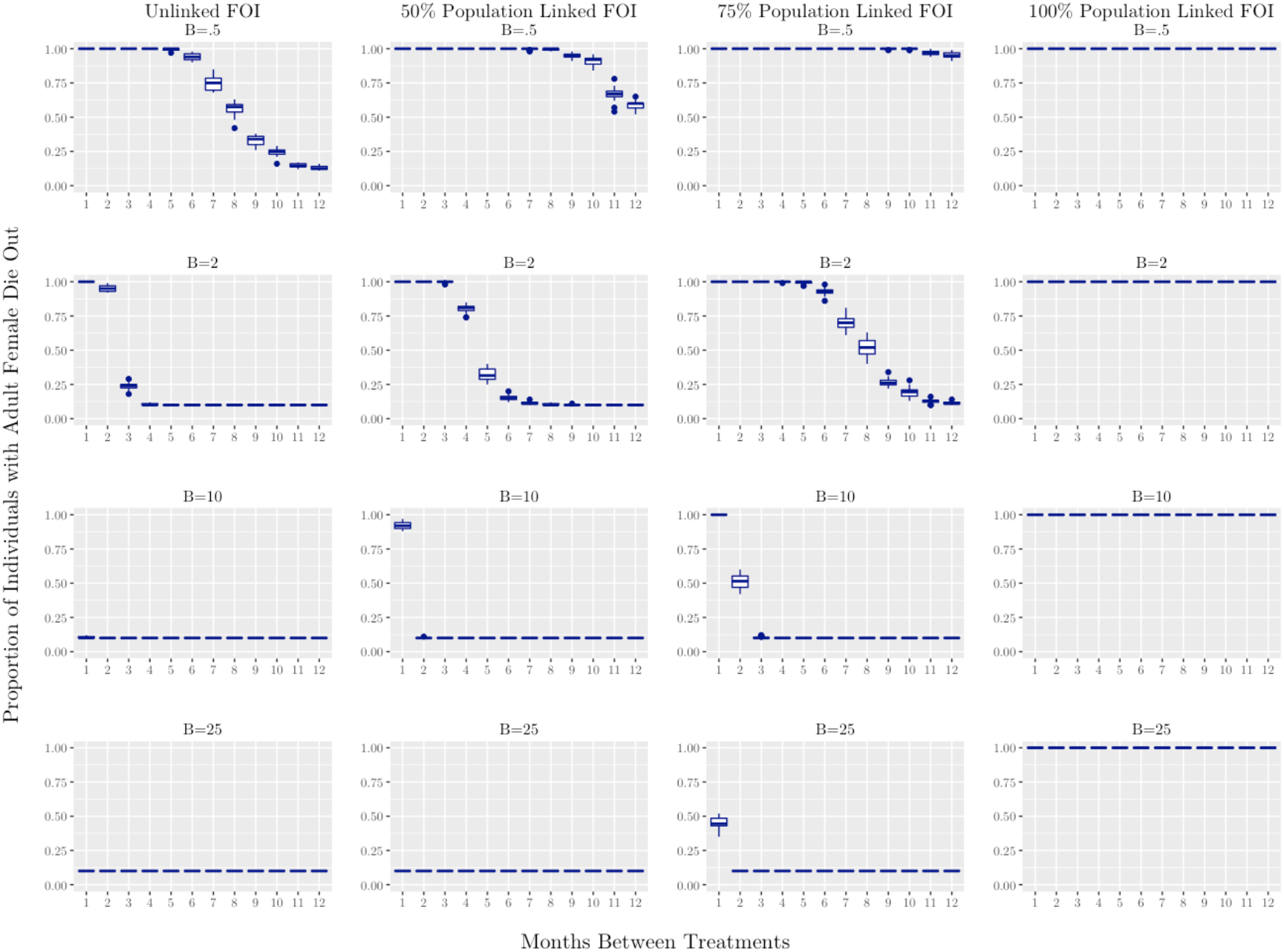
A measure of transmission interruption over 3-year intervention period under varying treatment timing regimes and FOI initial values (B) and linkage types. Large proportion of individuals with adult female schistosome die out is only possible under low FOI conditions or for moderate FOI with treatments every 1-3 months or higher coverage (75-100% linked FOI). High FOI regimes only achieve adult female die out with 100% linked FOI (complete population coverage) or high population coverage (75% linked FOI) and treatment every 1-2 months. The boxplots show the proportion of individuals who experience adult female die out over 1,000 runs.

The proportion of time without egg production, implying a break in transmission to the intermediate host, increases with decreasing time between treatments (Figure 7). In populations where treatment is applied to a broad spectrum of the population, the initial population burden has an influence on the time during which there is a break in transmission. proportion of time without contributing to transmission. Those populations with low initial burden have a greater proportion of time without egg production (SI Fig. 21). For situations with a moderate initial FOI (B=2), complete cessation of egg production is difficult to achieve and is only possible with broad treatment coverage or moderate coverage and treatment frequencies of 1-3 months (Figure 7). High initial FOI environments (B=10-25) sustain egg production no matter the frequency of treatment if there is not high population coverage (Figure 7). The proportion of intervention time with adult female and total schistosome die out mirrors these patterns and the individual-linked FOI showed a pattern similar to that of the 100% population linked FOI (SI Fig 23).

**Figure 7.**
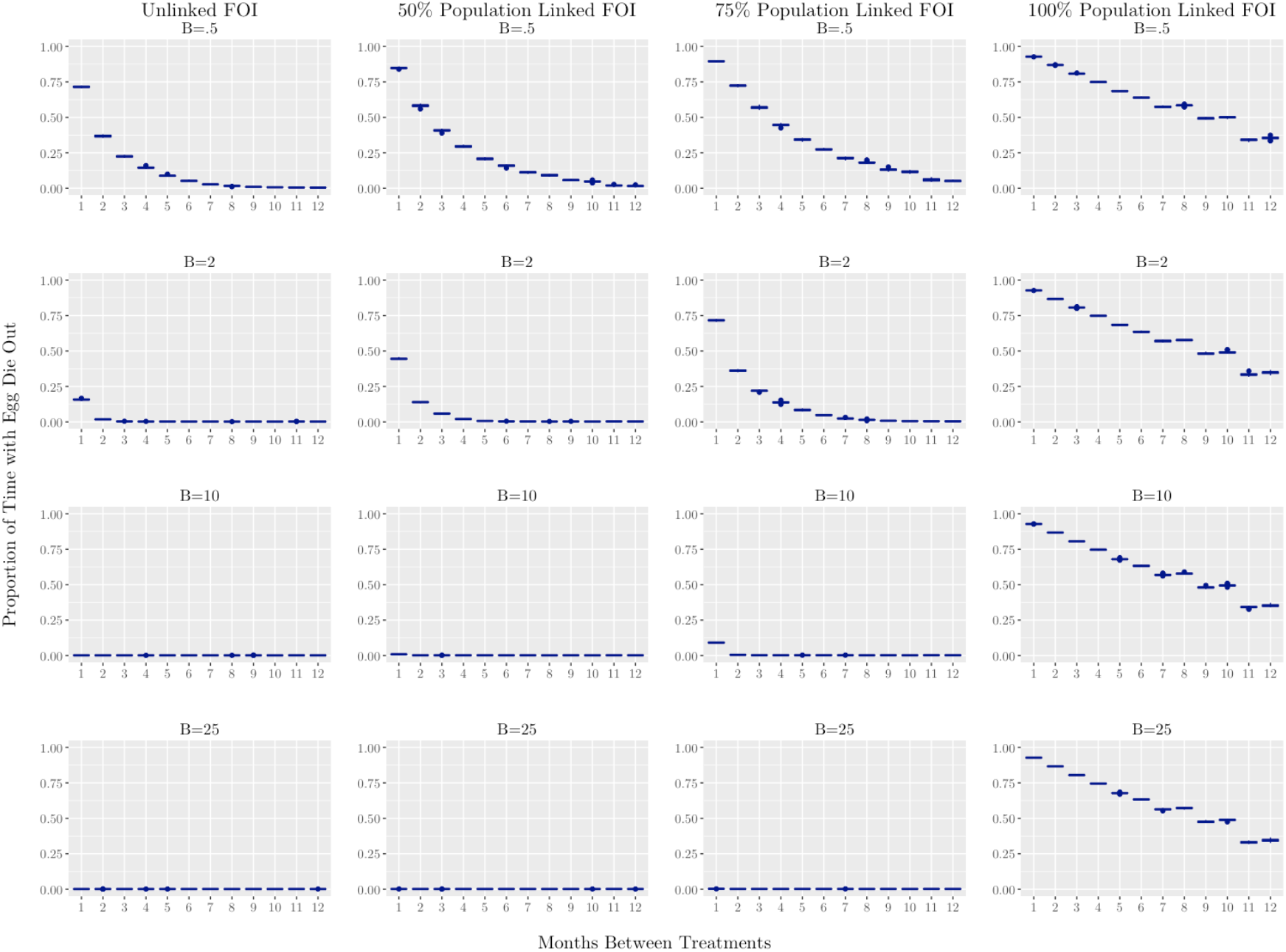
A measure of transmission interruption over 3-year intervention period under varying treatment timing regimes and FOI types and values. Substantial egg die out is only achieved in moderate to high (B=10-25) FOI levels if there is treatment bimonthly or feedback (scenario ii), with high population coverage (75-100% linked). The boxplots show the proportion of time during the intervention period when there were zero eggs produced with a high initial worm burden.

### Stage Dependent Efficacy

To demonstrate the impact of stage-specific schistosome mortality due to treatment with praziquantel we also consider two “control” models with no differential mortality due to MDA between schistosome age classes. These models employ identical praziquantel effectiveness for all age classes, at either a low (70% mortality) or standard (90% mortality) effectiveness with treatment (SI Figs. 2-3). Both the 70% and 90% praziquantel effectiveness have been found in field studies [20,27,30]. As expected, the 70% all-stage mortality displays greater overall total number of schistosomes after an intervention, though with large variance in MDA treatments at 11-or 12-month intervals (SI Figure 2). In general, the 90% all-stage mortality model has lower schistosome burden after an MDA intervention than is predicted by our stage-specific mortality model. With both all-stage mortality models, extremely frequent treatments, such as every 1-3 months, depress the total schistosome population levels more than we see in the stage-specific mortality model. This discrepancy can be attributed to the more rapid rebound in the adult schistosome population after treatment from the reservoir of juvenile stages that are more resistant to praziquantel. Therefore, a non-stage specific model may overestimate the effectiveness of a frequent praziquantel-based intervention.

An alternative drug, oxamniquine, primarily used in South America for the treatment of *S. mansoni*, was evaluated as it causes differing schistosome sex and stage mortality than praziquantel [17,31]. Of particular note, oxamniquine is more effective against juvenile schistosomes and adult males and less effective against adult female schistosomes than praziquantel [[32], SI Table 1]. The greater survival of adult females after oxamniquine treatment, coupled with the male skewed sex bias, leads to higher estimated schistosomes after a 3-year intervention than with praziquantel (SI Figs. 4-5). Similar to the all-stage mortality models, there are very low population sizes at extremely frequent treatment as oxamniquine effectively diminishes the juvenile schistosome population removing this potential reservoir (SI Figs. 4-5).

### Post – Intervention Populations

Reduction in mean worm burden is an important metric of the effectiveness of schistosomiasis control but the projected mean worm burden after the cessation of an intervention is an equally important outcome. Simulations of the rebound in adult mean worm burden two years post-intervention for the unlinked FOI model show approximately the level of infection expected with these average background FOI values in a situation pre-intervention (SI Fig. 1). The rebound trajectories show that under low and moderate FOI regimes, even those where the FOI is not linked to post-treatment egg burden, these populations may reach a new equilibrium and a lower burden regime (Figure 8). Under high FOI regimes, where there is rapid rebound in the schistosome population due to reinfection, there is still a large, if transient, reduction in burden. The model with the individual linked FOI mirrored the fully population linked FOI (SI Fig 24).

**Figure 8.**
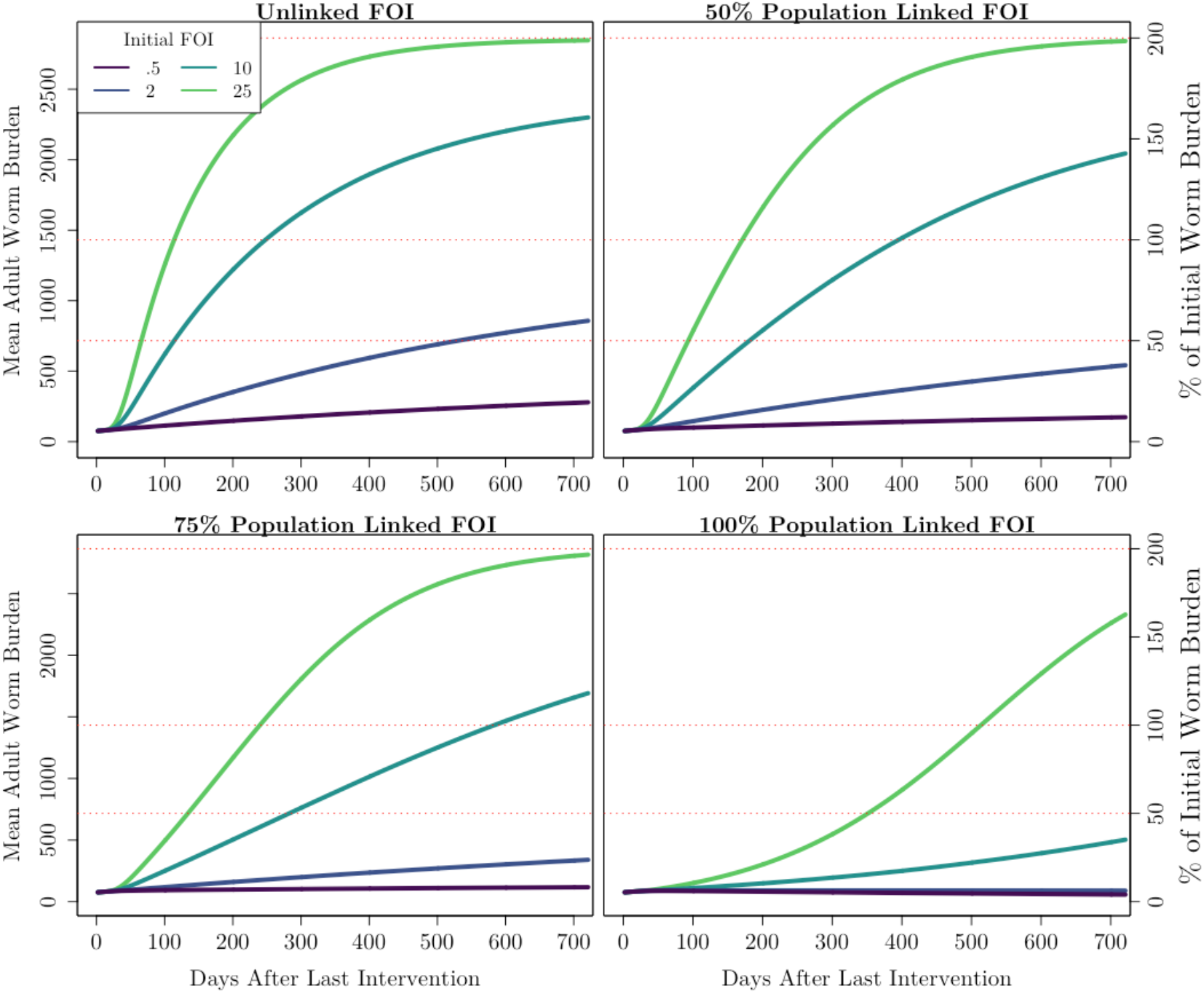
Schistosome population trajectory in 24 months after the cessation of 3-year intervention period where treatment was applied every 6 months. Under moderate and high FOI conditions there is rebound to pre-MDA schistosome levels within 2 years. The mean adult worm burden number under various FOI types and initial values (B) summarized over 1,000 runs. Dashed red lines indicate the mean worm burden equal to 50, 100, and 200 percent of the moderately infected population mean worm burden prior to the intervention.

## Discussion

Our results indicate that effective interventions are not one-size-fits-all. The severity of *S. mansoni* burden in the population prior to an MDA intervention, the proportion of the population that is included in the MDA and the impact on the FOI from those individuals not targeted in the MDA, are all important in determining effective intervention strategies. We propose that the variability in the success in MDA after multi-year intervention programs is in part due to the insufficient frequency (only once or twice a year) and the treatment of only the school-aged children segment of the population [22–24,33]. In multiple studies, school-aged children have been found to be disproportionately infected with *S. mansoni* and have been attributed to contribute to the bulk of transmission [22]. However, our results indicate that if they are not responsible for at least 75% of the force of infection to the intermediate snail host then the population as a whole will not reach a lower burden status quo. The importance of targeting a large proportion of the population for treatment has been noted in other recent modeling studies [7,8]. While elimination may not be possible without repeated treatment to at least 75% percent of the population at intervals shorter than the standard 12 months, it is important to note that the MDA at 12-month intervals does greatly reduce morbidity in the treated individuals. The reduction in morbidity, particularly in school-aged children is a primary short-term goal for these MDA programs [5]. The success of these interventions may be transient if the input of eggs into the aquatic environment is impacted, as is shown in the difference in the potential for rebound to pre-intervention burden levels and the time to rebound in these scenarios (Figure 8).

Rebound to pre -treatment schistosome population and population egg production is an important metric if the goal is elimination or a decrease in population burden to a lower endemic level. Rebound was rapid, occurring within two years under the majority of our scenarios. Our results show rebound under all but very low FOI conditions or low FOI and highly linked FOI, that is the repeated application of treatment to more than 75% of those contributing to transmission. This pattern is irrespective of longer treatment horizons. The stage dependent efficacy of the pharmaceuticals used impact the success of these MDA campaigns. The influence of reduced efficacy against juvenile parasites on the success of MDA campaigns in our study mirror the findings of a prior study; particularly in that better MDA outcomes over long time horizons and in high burden settings would be possible without this reduced efficacy in juveniles [11]. Our alternative drug option, oxamniquine, performed similarly to our “control” models with equal effectiveness across all schistosome life-stages by reducing the impact of a reservoir of juvenile life stages after treatment. However, though this did slightly alter the speed of rebound to pre – treatment burden levels, the rebound to prior burden levels did still occur under the same FOI and treatment frequency regimes. In the absence of a more efficacious drug treatment option, which would be expected to more readily achieve elimination, efforts should be focused on breadth of treatment in the population and more frequent treatment [11]. Without elimination or transition to a lower endemic burden level under moderate or high FOI conditions the impact on long-term population burden is often transient due to rebound.

Our study also explores scenarios with initial FOI and worm burdens which do not resemble the predicted long-term trends of our model without treatment (SI Figure 1). While there is a potential mismatch between the burden level and the assumed initial FOI there are specific circumstances in which the FOI may not correspond to the equilibrium level of prevalence expected for that level of infection risk. For instance, a high initial population burden and a low level of infection risk (FOI) could be the result of other newly introduced intervention measures such as infrastructure improvements, molluscicide applications or a snail removal effort. There is also a potential mismatch between a low initial population burden and a high FOI, this could occur in an instance where the estimates of population infection are based on a non-representative sample of the population with a lower-than-average burden, or where there is a rise in the FOI due to alteration of exposure. An escalation in risk of infection may be due to increases in the snail host population, an outside influx of infection, or an alteration of the water sources used by the population. The impact of the worm burden distribution in the population between those individuals targeted by MDA and those who do not undergo regular treatment may be better understood by future work to validate this model through the expansion of prevalence surveys and MDA to ascertain the population level burden and the background FOI prior to and after MDA campaigns.

There are several limitations of this study, notably the simplification of the relationship between egg production and force of infection, as we did not explicitly model the intermediate snail host population. This simplification was included to make the model generalizable to populations with varying *S. mansoni* burden levels. Future work could parameterize the model to include the proportions of populations treated with MDA and calibrate the model to the pre-intervention and post-intervention burden levels in the population. Also, this model assumes a consistent relationship between population egg output and risk of infection. The risk of infection may be altered by seasonal fluctuations in the intermediate snail host population or human water source exposure. Though widespread praziquantel resistance has not been identified, and in some cases there was robust diversity even after MDA, there is the possibility of developing praziquantel resistance with increased selection pressure under more frequent and expansive treatment regimes [34,35]. Resistance to praziquantel could drastically alter the efficacy of praziquantel and the success of MDA campaigns. The possibility of acquired immunity to *S*.*mansoni* infection has been debated, and prior modeling studies have shown that over long time horizons it may in fact hinder the impact of MDA, as we are primarily focused on shorter time horizons we have not included acquired immunity in our model [36,37].

As has been mentioned in other work, due to the robustness of the Schistosoma system and the possibility of reinfection, a more comprehensive intervention plan is needed for elimination or long-term reduction in high burden situations [21,26,38–41]. Our study suggests that there are no substantial benefits for more than six treatments a year under most situations, and in the majority of our scenarios there are diminishing returns after three treatments a year. This stagnation in effectiveness concurs with recent studies which posit that the expansion of treatment to include adults and more frequent treatment timing is necessary to reach elimination and that the addition of snail control would be particularly helpful in high-risk communities [19,21,38,39]. Another potential avenue for combined intervention, vaccines, are under development but are not currently a viable strategy for control, though in the future may be a powerful tool in conjunction with MDA and other intervention strategies [41].

However, we also find persistence in low and moderate FOI and burden populations when 50% or less of the population contributing to transmission is treated with praziquantel. This persistence is due to reinfection, as the FOI is not substantially altered, and the survival of the juvenile schistosomes after praziquantel treatment. Therefore, if logistics and resources allow for investment in more frequent treatment, greater treatment coverage, or longer intervention campaigns, our results indicate that two or three treatments a year are advantageous, but to achieve elimination very broad community coverage is necessary. Furthermore, interventions longer than a few years are not substantively more effective than shorter interventions of the same coverage and frequency.

## Data Availability

The data that support the findings of this study, in the form of R code, are publicly available from the following Github repository:https://github.com/larissaanderson/Within-Host-S.-Mansoni-Modeling-Code

